# A community-based participatory research approach to the development, refinement, and distribution of Test-to-PrEP: A peer-to-peer distributed at-home HIV self-test and prevention information kit in Miami-Dade, Florida

**DOI:** 10.1101/2023.12.22.23300158

**Authors:** Stefani Butts, Edda Rodriguez, Lacey Craker, Ariana L. Johnson, Patrick Whiteside, Jakisha Blackmon, Sonjia Kenya, Mariano Kanamori, Susanne Doblecki-Lewis

## Abstract

In the US, there is a need for interventions that address gaps in awareness, interest, and uptake of HIV testing and biomedical HIV prevention strategies, such as pre-exposure prophylaxis and non-occupational post-exposure prophylaxis. The Test-to-PrEP intervention; an HIV self-test bundled with prevention information that was distributed via a social network strategy, was found to be effective at bridging said gaps. This manuscript presents the development and design of Test-to-PrEP, in which a community-based participatory research approach was used.

The intervention combines peer-to-peer distribution of HIV self-testing kits with tailored HIV prevention education. Key features include culturally sensitive educational materials, a strategic emphasis on the connection between intervention distributors and recipients, and rigorous training protocols.

Our process led to the creation of materials that were neutral to sexual identity, highlighting the importance of inclusivity and cultural relevance. It also led to a mechanism that allows enhanced network member selection. Stakeholder guidance ensured the initiative was well-aligned with community needs, enhancing its potential acceptability and effectiveness.

By centering community needs and cultural nuances, interventions like Test-to-PrEP can potentially increase their reach and efficacy. Our development process underscores the importance of community engagement, cultural relevance, and well-defined reporting.

## Introduction

In the United States (U.S.), nearly 1 in 7 individuals living with human immunodeficiency virus (HIV) is unaware of their status (Centers for Disease Control and Prevention, 2021a). As such, there is a critical need to address current gaps in awareness, interest, and uptake of HIV testing and biomedical HIV prevention strategies, such as pre-exposure prophylaxis (PrEP) and non-occupational post-exposure prophylaxis (nPEP), particularly among communities disproportionately impacted by HIV.

Strategies such as home-based HIV testing kits, peer-led education interventions, and mobile clinics may be more approachable and acceptable compared to traditional clinic-based HIV prevention services (Hillis, Germain, Hope, McVeigh, & Van Hout, 2020; Rousseau, Julies, Madubela, & Kassim, 2021). Social network strategies to disseminate HIV testing and prevention efforts through peer networks (e.g., sexual and friendship) can further enhance reach. The combination of such strategies presents an opportunity to de-medicalize and simplify HIV prevention services and can reduce barriers that hinder access to HIV testing and biomedical HIV prevention tools (Chimbindi & Shahmanesh, 2022; Hillis et al., 2020; Rousseau et al., 2021).

Miami-Dade County, Florida (MDC) has among the highest HIV incidence and prevalence rates in the nation (Centers for Disease Control and Prevention, 2022). In MDC, nearly 14% of those living with HIV are unaware of their HIV status (U.S. Health and Human Services, 2019). As of 2018, the most recent year with available PrEP coverage data, less than 20% of individuals living in MDC with indications for PrEP had received a prescription (U.S. Health and Human Services, 2019). Together these indicators highlight the need for increased efforts to improve HIV testing and linkage to biomedical prevention services in MDC.

Designing strategies to increase priority populations’ knowledge of, access to, and use of HIV testing and prevention strategies requires an insider’s understanding of what is acceptable and feasible for each community. Community-based participatory research (CBPR), which emphasizes co-learning, the sharing of expertise, and distributing decision-making power among community members, organizational representatives, and academic researchers (Coughlin, Smith, & Fernandez, 2017; Israel, Eng, Parker, & Schulz, 2012), is an effective way to work with communities to gain this inside perspective (Coughlin et al., 2017; Israel et al., 2012). CBPR also allows researchers to identify and develop strategies in partnership with the community, increasing the intervention’s reach, acceptability, appropriateness and ultimately, improving testing and prevention outcomes (Coughlin et al., 2017; Israel et al., 2012).

Consistent with this approach, we employed an iterative CBPR model that builds on the work of Rhodes et al. (2013), to conceptualize, design, and disseminate Test-to-PrEP. This intervention leverages peer networks to distribute bundled HIV self-tests, along with HIV, PrEP and nPEP information. The objective of this report is to delineate the Test-to-PrEP framework (Table 1), outlining its development, lessons learned, and potential directions for scaling and sustaining this community-focused intervention.

**Table 1.**
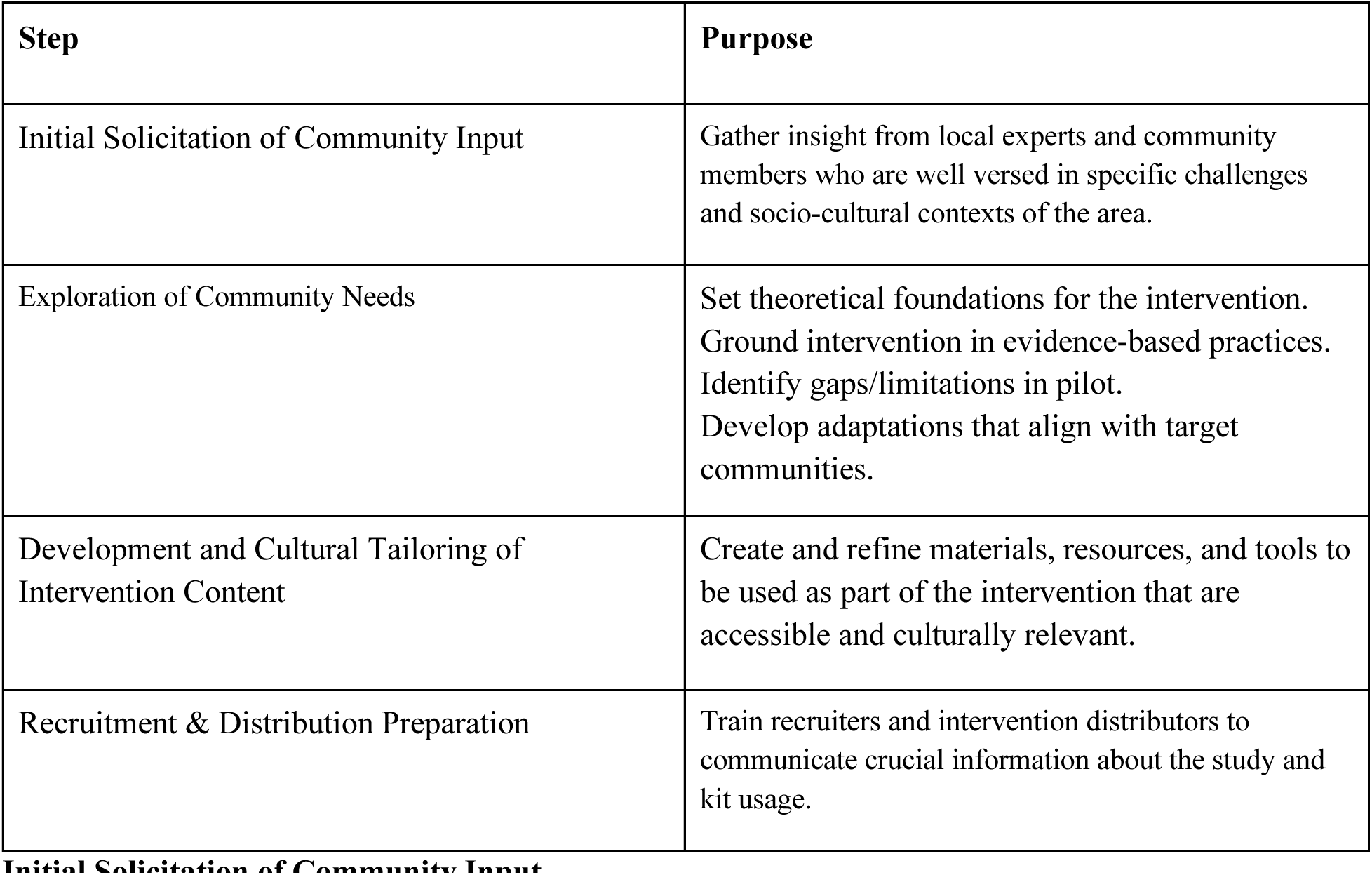
Test-to-PrEP Iterative Framework

## Materials and Methods

### Setting

The Rapid Access Wellness (fixed clinic) and Mobile PrEP initiatives, at the University of Miami, are health care clinics that provide sexual health services including STI/HIV testing, STI treatment, PrEP, nPEP, and rapid entry into HIV care at no cost to clients. The majority of clients served by these clinics are Hispanic/Latino, immigrant, men-who-have-sex-with-men (MSM). The clinic team collaborates regularly with both academic and community-based partners to create and deliver evidenced-based and community-prioritized health care services.

### Intervention Concept Development

The “Test-to-PrEP” intervention builds upon a pilot study which engaged existing PrEP clients to distribute HIV self-tests to members of their social networks (King et al., 2022). The intervention was proposed in response to qualitative feedback from pilot study participants supporting the expansion of the pilot and inclusion of educational materials regarding PrEP and nPEP (King et al., 2022). To ensure that “Test-to-PrEP” would be coherent with the needs and priorities of the community, a CBPR-based approach was used for its development.

### Initial Solicitation of Community Input

In April 2020, a 10-member community advisory panel was assembled. Panelists included the director and staff members from Prevention305; a community-based non-profit HIV prevention organization, the director and staff members from CHAMP; a community service program that provides HIV testing, linkage to care, and advocacy to individuals from Miami’s diverse Black neighborhoods, and PhD and masters-level community health experts in the HIV prevention field from UM’s Social Networks and Latino Health Disparities lab. Panelists were Miami-Dade residents, individuals identifying as Hispanic/Latino MSM, immigrants, Black cisgender women, and/or individuals experienced in HIV testing and PrEP engagement within Miami-Dade County. The group met weekly to discuss content and practical strategies for development. Meeting notes were recorded in real time and reviewed regularly by the study team.

### Exploration of Community Needs

This stage was conducted using an iterative approach, garnering input and feedback through discussion of current HIV-testing practices and perceived barriers and facilitators of HIV testing and HIV self-test (HIVST) program implementation. Panelists were encouraged to consider this data in the context of their experiences.

### Review of Foundational Work

First, qualitative data collected and analyzed during the pilot study (King et al., 2022) were presented to the panel to identify opportunities for feasible and swift adaptations. Four broad potential improvement areas were found, these included: intervention-distribution logistics, data collection on intervention recipients/users, HIV prevention education for intervention recipients/users, and training on HIVST-handling, use, and distribution, for intervention distributors. Each area identified was subsequently divided into more specific subcategories, allowing for a nuanced understanding and targeted approach to addressing the individual challenges within each domain.

### Literature Review

Panelists then reviewed literature on existing U.S.-based HIV self-testing interventions that leveraged social network strategies (SNS) for distribution, examining their potential for scale-up, sustainability, and integration into the Test-to-PrEP intervention. At the time of this review, only two studies that fit location and methodology criteria had been published (Lightfoot et al, 2018 & Wesolowski et al, 2019). Panelists characterized each study by relevant similarities and differences.

Both studies found that SNS strategies were effective at increasing testing among MSM, particularly MSM of color who had never tested for HIV (Lightfoot et al, 2018 & Wesolowski et al, 2019). However, while the study by Lightfoot and colleagues recruited intervention distributors who were living with and without HIV and provided training on HIVST use (Lightfoot et al, 2018), the Wesolowski group recruited distributors who were living with HIV and did not train participants on use of the self-test (Wesolowski et al, 2019). The Lightfoot study also implemented an electronic mechanism to collect data from HIVST users/recipients (Lightfoot et al, 2018). Both studies recruited HIVST distributors mainly through social media and dating applications and focused on recruiting distributors who were Black and Hispanic MSM (Lightfoot et al, 2018 & Wesolowski et al, 2019). Neither study recruited cisgender women for distribution or included HIV-prevention educational materials with the HIVST.

Intervention adaptations to address the gaps found within the pilot were then identified. Gaps and proposed adaptations are summarized in Table 2.

**Table 2.** Summary of Intervention Optimization: Opportunities for Enhancement and Proposed Adaptations

| **Opportunity** | **Adaptation** |
| --- | --- |
| **Kit Distribution Logistics** |  |
| Underrepresentation of Black MSM and cisgender women among kit recipients | Focused recruitment of Black and cisgender female PrEP clients |
|  | Develop inclusive intervention materials & messaging |
| Lack of confidentiality during HIVST distribution due to packaging of materials | Develop “discrete” intervention packaging |
| Unstructured Test-to-PrEP distribution | Survey-facilitated creation of distribution plan |
| Limited intervention reach | Allow recipients to become distributors |
| **Data Collection on Intervention Recipients/Users** |  |
| Secondary reporting of intervention users/recipients’ HIV prevention knowledge and HIVST use | Create and disseminate user/recipient data collection tool |
| **HIV Prevention Education for Users/Recipients** |  |
| Lack of prevention information materials accompanying the HIVST | Develop prevention-education materials to be included within the intervention |
| **Recruitment & Distribution Preparation** |  |
| Limited knowledge on handling & usage of HIVST among distributors | Develop and conduct formal training with intervention-distributors |

### Outcomes

Development and Cultural Tailoring of Educational Materials and Data Collection Tool Guided by findings and insights from the panel, the first drafts of intervention content and data collection tools were formulated. Materials included a palm-sized card with a quick response (QR) code that opened the survey and instructions for taking it when scanned, a six-page booklet that detailed information about HIV, PrEP, and nPEP and an opaque tote bag with RAW/mobile PrEP names and logos printed on the front for transporting the HIVST and educational materials.

### Educational Booklet

Information included within the booklet was based on priorities expressed by pilot study participants and guidelines from the World Health Organization (WHO) for implementing HIV self-testing programs (King et al., 2022;WHO, 2021). This resource was divided into two sections. The first section focused on providing foundational knowledge about HIV. This included considerations for before using the HIVST, related to recency of exposure, guidance on interpreting test results, and actionable steps depending on the test’s outcome. The second section described PrEP and nPEP HIV prevention strategies. Additionally, the booklet offered clinic referral details and provided the process for scheduling appointments.

Pictures of reactive and non-reactive HIVST results were included in the booklet to facilitate comprehension among intervention recipients. The use of pictures in health communications can improve attention, recall, and adherence to instructions and can be particularly helpful for individuals with low literacy skills or limited language proficiency (Houts et al., 2006).

Qualitative feedback garnered from the pilot study (King et al., 2022) and recommendations from panelists supported a design that was gender- and sexual identity-neutral, leading to inclusion of photos and language within the booklet that were not anchored to any specific gender or sexual orientation. It was believed that this would facilitate broader applicability and resonance with a diverse range of participants. Indeed, results from a study that assessed perceptions of HIV testing campaigns among Black and Latino individuals found that these groups were more likely to find messaging acceptable if they were not focused on the LGBTQIA+ community (Drumhiller et al., 2018).

### Survey & Data Collection Design

Survey questions were developed based on priority areas identified during the “Exploration” step. They encompassed a broad range of topics, from user demographics and HIVST outcomes to patterns of HIV risk behaviors. Likert-scale items queried HIVST-users’ understanding of HIV, self-testing processes, and preventive measures. Distinctive features of the survey were its emphasis on relationship dynamics between intervention recipients/users and distributors and that it asked recipients/users to rate their pre- and post-intervention PrEP & nPEP knowledge, evaluate the intervention-information’s quality, and express their propensity towards future PrEP use. The survey assessed primary outcomes of the larger study, which are reported elsewhere (Johnson et al., 2023).

It was observed that prior HIVST interventions recruited distributors for whom limited background knowledge was known and did not possess reliable mechanisms to associate these distributors with HIVST recipients (Lightfoot et al, 2018 & Wesolowski et al, 2019). Panelists recognized this as an opportunity to leverage the comprehensive demographic and sexual risk data on distributors, already collected by RAW/mobile PrEP. They hypothesized that this data-rich context would facilitate a more in-depth characterization of effective intervention-distributors. To establish a direct connection between intervention recipients and respective distributors, each QR code was embedded with an individual identifier that linked distributor-recipient pairs automatically when scanned, further enhancing the traceability and potential feedback mechanisms of the Test-to-PrEP intervention.

### Language Adaptation

To ensure inclusivity and broad accessibility, all materials and the survey were translated into Spanish. Spanish versions were then circulated among native Spanish-speaking panel members and project staff. This was done to ensure linguistic accuracy and cultural relevance, as research has shown that social context and local specificity of language is important for encouraging acceptability of public health messages directed at specific communities (Fields et al., 2020)

### Pretesting & Refining Materials

The drafted Test-to-PrEP intervention concept was then shared with a CBPR and Social Networks graduate class at the University of Miami, acting as a proxy focus group. Their feedback, combined with insights from current PrEP program clients, led to essential revisions, particularly concerning privacy and clarity. As an example, participants expressed concerns about privacy, which led the panel to select a material for the tote bag that was opaque. Their feedback also influenced the information that was displayed on the outside of the bag; logos and images used did not explicitly indicate HIV related content or describe services provided by the clinics.

**Figure 1.**
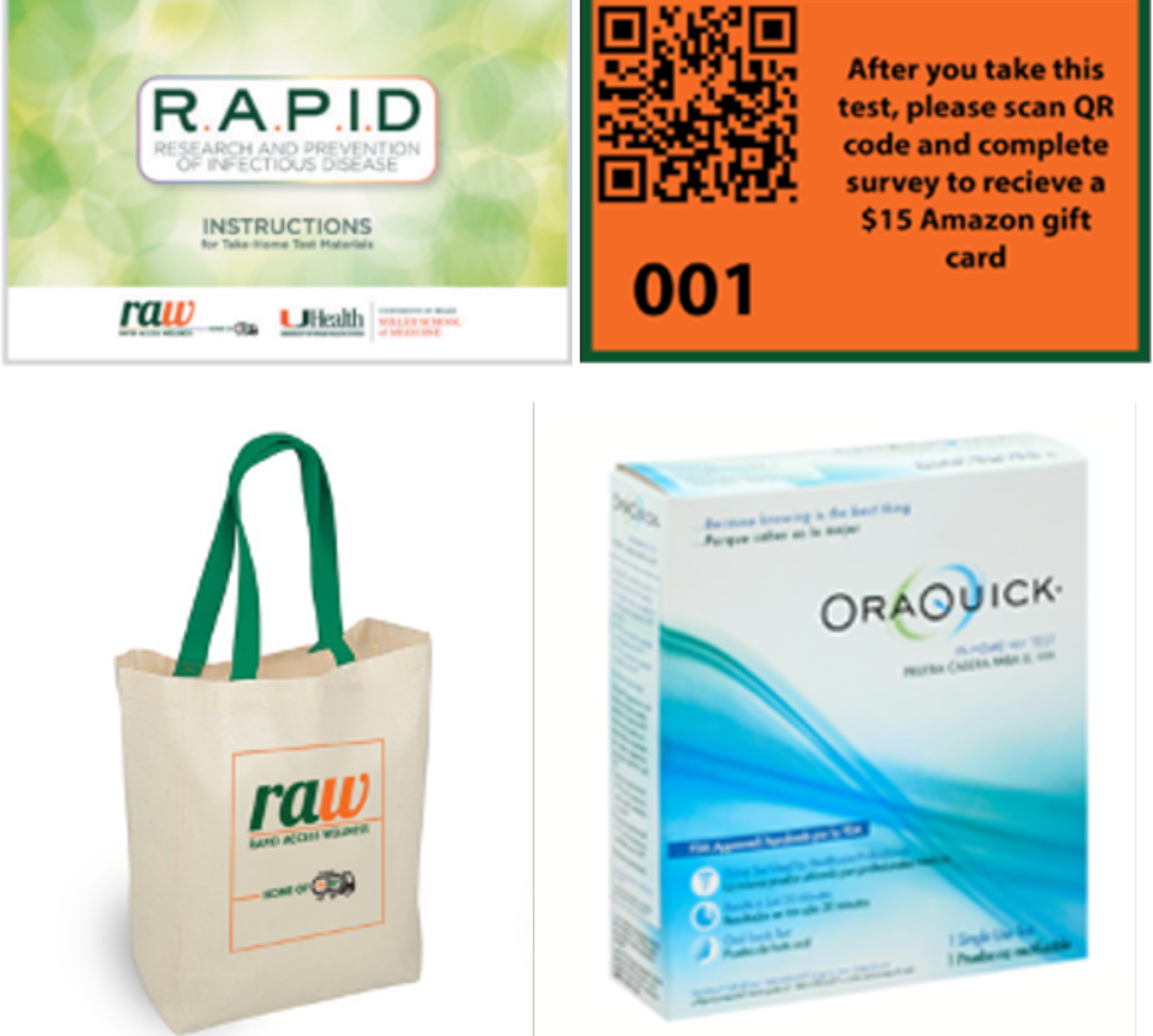
Materials Visualization

### Recruitment & Distribution Preparation

Aiming to equip study recruiters and intervention distributors with adequate knowledge on Test-to-PrEP content and HIVST procedures, panelists developed a Test-to-PrEP “training module” for study recruiters. The module was based on suggestions described within pilot study data, related to distributor preparedness and self-efficacy (King et al., 2022), and grounded in the “Train-the-Trainer” approach. Train-the-Trainer is a capacity and dissemination multiplier model in which subject matter experts train others to become trainers themselves (Yarber et al., 2015). It allows sharing of knowledge and skills with a wider audience and helps develop training expertise amongst said audience (Yarber et al., 2015).

The module included role-play scenarios, helping recruiters familiarize themselves with potential real-world interactions with PrEP clients. An accompanying brief document with key bullet points was also created for quick reference. This document included short prompts that reminded recruiters to emphasize the most important information about Test-to-PrEP when training distributors. Two outreach coordinators, possessing relevant clinical, research, and linguistic competencies, underwent intensive two-day training on this module.

## Discussion

We developed the Test-to-PrEP intervention, which strategically integrates peer-to-peer distribution of HIVST with tailored HIV prevention education, through a robust CBPR approach. With the understanding that interventions often have increased efficacy when tied to community needs and assets, our process of design and dissemination of a community-centered intervention that aims to increase HIV testing and prevention knowledge may be applicable to development of similar interventions.

Central to the success of the Test-to-PrEP development process was the engagement of community stakeholders and organizations. Such engagement ensures alignment with specific experiences and preferences of priority populations and emphasizes the responsive and dynamic nature of the initiative, as evidenced by findings from the pilot study and other interventions (King et al.,2022; Lightfoot et al, 2018; Wesolowski et al, 2019) and bolstered by real-time feedback. CBPR research methods have been shown to enhance the acceptability of health interventions by involving community members at every stage, from design to evaluation (Viswanathan et al., 2004), and employing these methods has been linked to the creation of health interventions that are more sustainable and equitable (Viswanathan et al., 2004).

After deployment of the Test-to-PrEP kit, intervention recipients/users indicated the high quality of information provided (Johnson et al., 2023). This aligns with prior research suggesting that the efficacy of information-based interventions hinges on the caliber of the information presented, the modality of its delivery, and the specific characteristics of the target audience (Gagliardi & Brouwers, 2015). The importance of using high-quality information for these interventions plays a crucial role in equipping individuals with the insights necessary to comprehend their health risks and benefits, instigate behavioral change, and make informed decisions about their health (Epping-Jordan et al., 2004).

An important feature of Test-to-PrEP was the purposeful tailoring of language, imagery, and content to resonate across diverse genders, sexual orientations, and cultural backgrounds, as our objective was to amplify the program’s reach and enhance its efficacy. This aligns with the observations of Drumhiller et al. (2018), who ascertained that HIV prevention messages garnered more public acceptance when they highlighted general risk factors for HIV and refrained from using terms like “MSM” or explicit depictions of sexual activity. Further, Test-to-PrEP was deliberately inclusive of cisgender women as intended recipients as this group was perceived to have been left out of many HIV prevention initiatives.

## Conclusion

We present a CBPR-based process for development of Test-to-PrEP, an intervention to increase HIV testing and PrEP information. We hope that our description of the Test-to-PrEP process may aid others in development of similar interventions, and that the inclusion of this process in intervention development will result in amplification of the potential health impact.

Future research will assess the efficacy and implementation needs of Test-to-PrEP deployment. Periodic reevaluation and adaptation of the model in partnership with community stakeholders is also planned, to ensure that our strategy remains relevant and aligned with the evolving needs of diverse communities.

## Data Availability

This is a study protocol describing a community-based participatory method of intervention development. Hence, no data are associated with this manuscript

## Acknowledgements

This work was supported by Miami Center for AIDS Research Ending the HIV Epidemic administrative supplement (NIH/NIAIDS P30AI169643) awarded to M.K. and S.D.L

## Author Contributions

SDL and MK conceived the idea presented. SB, ALJ, ER, and LC wrote the manuscript with support from SDL and MK. SDL and SB supervised the project. All authors were involved in the intervention’s iterative planning and development process, its revision for intellectual content, and approved the final version of the manuscript. All authors agree to be accountable for all aspects of the work.

